# US County-level Structural Racism Effect Index and Cardiovascular Disease Mortality among Older Adults: A Bayesian Spatiotemporal Modeling

**DOI:** 10.64898/2026.07.10.26357792

**Authors:** Tahmina Begum, Md. Shahjahan, Hrishikesh Chakroborty

## Abstract

**Background:** Cardiovascular disease (CVD) remains the leading cause of mortality among older U.S. adults, yet the contribution of neighborhood-based structural racism remains inadequately quantified. This study quantifies the association between the Structural Racism Effect Index (SREI) and CVD mortality among adults aged ≥65 years, evaluating how this relationship varies across U.S. geographic regions to identify key areas for intervention.

**Methods:** This ecological study applied a hierarchical Bayesian spatiotemporal framework to 2017–2020 Centers for Disease Control and Prevention (CDC) Wide-Ranging Online Data for Epidemiologic Research (WONDER) data to estimate the association between SREI and CVD mortality across 3,007 U.S. counties. SREI was modeled continuously and categorically, adjusting for sociodemographic covariates. Population attributable fractions (PAF) and attributable deaths (AD) quantified the potentially preventable burden and its spatial disparities.

**Results:** From 2017 to 2020, approximately 2.79 million CVD deaths were observed, with significant spatial clustering (Moran’s I = 0.35, *p* < 0.001). Each standard-deviation increase in SREI was associated with 13% higher CVD mortality (IRR: 1.13, 95% CrI: 1.12–1.15). A positive dose-response gradient was observed across SREI quartiles, with mortality 24% higher in the highest quartile than in the lowest (IRR: 1.24, 95% CrI: 1.20–1.28). The PAF was 6.94% (95% CrI: 6.13–7.73), corresponding to 193,472 potentially preventable deaths. High exceedance probabilities (>0.95) were concentrated in the Southeast, Appalachia, and the Midwest.

**Conclusions:** Structural racism is a spatially patterned, dose-dependent predictor of older adult CVD mortality, underscoring the need for public health monitoring and neighborhood-based upstream interventions where disease burden is concentrated.

## INTRODUCTION

Cardiovascular disease (CVD) remains the leading cause of morbidity and mortality in the US, accounting for one in three deaths, with the age-adjusted rate of 218.3 per 100,000 in 2023.^1^ This burden disproportionately affects older adults, who represent 80% of CVD-related fatalities.^2^ While traditional clinical (such as hypertension, diabetes, hypercholesterolemia, and obesity) and behavioral factors are primary modifiable drivers,^3^ socioenvironmental determinants further constrain the outcome by restricting access to healthcare and health-promoting resources.^4,5^ Despite medical advances, persistent regional disparities necessitated geographic risk analysis to inform targeted intervention.^6^ These neighbourhood-disparities interact intricately, and often synergistically and bidirectionally.^7^

Neighbourhood structural racism—manifested through residential segregation, disinvestment, and resource maldistribution—serves as a fundamental barrier to health equity and drives profound mortality inequities.^8–11^ Capturing these compounding effects requires granular, multi-domain measurement of neighborhood structural disadvantage, essential to inform targeted public health interventions.^12^ Yet research linking such indices, including the Structural Racism Effect Index (SREI), to CVD mortality, among high-burden older adults, remains limited. Existing studies focus predominantly on individual-level demographics and clinical risk factors,^13,14^ leaving county-level geographic disparities unexplained.^15^ While prior research demonstrates that cardiovascular health declines as SREI scores rise,^16,17^ how county-level structural disadvantage shapes CVD mortality among older US adults and its spatial heterogeneity remains unaddressed. Characterizing these patterns requires geographically explicit methods.^18^ To our knowledge, no prior study has applied spatiotemporal disease-mapping models to assess multi-domain structural racism and county-level CVD mortality in this demographic.

We address this gap by applying a hierarchical Bayesian spatiotemporal framework with county-level covariate adjustment to examine the relationship between SREI and CVD mortality among US adults ≥65 years. We hypothesize that higher structural disadvantage predicts elevated CVD mortality, and a substantial proportion of the attributable burden is reducible through population-level shifts in structural conditions. This analysis provides a national assessment of how embedded social structures translate into preventable, fatal CVD outcomes.

## METHOD

### Data source

This study investigated structural neighbourhood factors and CVD mortality among U.S. adults aged ≥65 years. County-level CVD mortality (2017–2020) was obtained from the Centers for Disease Control and Prevention Wide-Ranging Online Data for Epidemiologic Research (CDC WONDER) database, which captures underlying causes of death across all 50 states and the District of Columbia; counts <10 were suppressed.^19^ The primary exposure, place-based structural disadvantages, was operationalized using the Structural Racism Effect Index (SREI) – a validated, nine-domain census tract-level composite, developed by Dyer and colleagues (2023) using the U.S. Census Bureau’s American Community Survey (ACS) 2015–2019 five-year estimates.^20,21^ Sociodemographic indicators were derived from the ACS database via the Census application programming interface (API) to ensure temporal alignment.^21^ County-level covariates included 2019 Supplemental Nutrition Assistance Program (SNAP) participation from the US Department of Agriculture (USDA) Economic Research Service (ERS) Food Access Research Atlas,^22^ and the 2018/2020 Social Vulnerability Index (SVI) from the Agency for Toxic Substances and Disease Registry (ATSDR) and the CDC, which integrates sixteen subcomponents across four domains (socioeconomic status, household composition/disability, minority status/language, and housing/transportation).^23^ Five datasets were merged by county’s Federal Information Processing Standard (FIPS) codes, resulting in 3,010 initial counties; excluding 1 county for missing mortality and 2 counties for missing SREI score, left 3,007 final analytical sample. Nine island counties were retained using a zero-policy constraint in spatial modeling to account for their lack of geographic neighbors and to avoid selection bias. Analyses ran from February to May 2026. All data were publicly available and de-identified; the study was exempt from Institutional Review Board approval and followed Strengthening the Reporting of Observational Studies in Epidemiology (STROBE) guidelines.^24^

### Assignment of exposure

The SREI is a validated composite score that spans nine domains: built environment, criminal justice, education, employment, housing, income and poverty, social cohesion, transportation, and wealth, constructed from multiple data sources (Table S1).^20^ Within each domain, geographic indicators were standardized, summed into domain-specific scores, and rescaled for cross-domain comparability.^20^ The final SREI score represents the mean of these standardized domain scores, with higher values indicating greater structural disadvantages.^20^ Tract-level scores were aggregated to the county-level using a population-weighted mean for proportional representation of populous areas. SREI was modeled continuously (z-score) and using distribution-based quartiles (Q1–Q4), where Q4 represented the greatest structural racism.

### Assignment of Outcome

CVD deaths were identified from death certificates using ICD-10 codes: hypertensive heart diseases (I10-I15), ischemic heart diseases (I20-I25), other heart disease (I30-I51), cerebrovascular diseases (I60-I69), diseases of arteries, arterioles, and capillaries (I70-I78), and other circulatory disorders (I95-I99). Deaths were aggregated over 2017-2020 to stabilize the estimate stability; counts <10 deaths were suppressed.^19^ Age-adjusted mortality rates (AAMR) per 100,000 were calculated via direct standardization to the 2000 US standard population.^25^ Expected deaths were calculated using CDC WONDER population estimates.

### Covariates

To control for potential confounding based on established associations, we adjusted for a *priori* county-level covariates.^17^ ACS demographics included self-reported biological sex (male, female), race, and ethnicity (white, Black, Asian, and Hispanic). Socioeconomic indicators comprised proportions of married individuals, bachelor’s degree holders, median household income, persons with disability, homeowners, and urban-rural status. SVI (continuous percentile ranking, 0-1) and SNAP participation (continuous measure) were included to capture broader vulnerability and food access barriers.

## STATISTICAL ANALYSIS

### Diagnostics, Descriptive, and Spatial Analytic Statistics

Statistical analyses were conducted in RStudio (v4.5.3). County identifiers were cross-checked to ensure accurate data linkage. Variable distributions, outliers, and transformation needs were assessed using histograms, density plots, and boxplots. SREI was standardized (z-score), and multicollinearity was assessed using correlation matrices and variance inflation factors. County characteristics were presented by SREI quartiles, summarizing continuous variables as medians (Interquartile ranges, IQR) and categorical variables as frequencies (%). Spatial exploratory analyses included county adjacency weights to evaluate spatial autocorrelation via Moran’s I. Choropleth maps and boxplots examining AAMR and expected deaths across SREI quartiles. Descriptive statistics for CVD mortality were stratified by SREI quartiles.

### Bayesian spatiotemporal modeling

A hierarchical Bayesian spatiotemporal negative binomial model was applied to account for overdispersion and estimate the association between SREI and county-level CVD mortality, with county population as an offset:

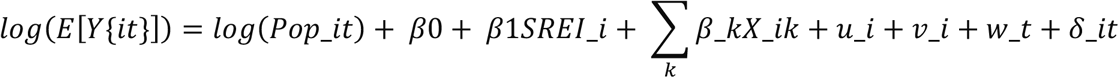

Here, [*Y*{*it*}] the CVD deaths in the county i during year t; log(*Pop*_*it*) the county-specific older adult population offset in county i at each time t. *β*0 is the intercept, *β*1 is the SERI fixed-effect coefficient, and *X*_*ik* represent county-level covariates. Spatial random effects were modeled using the Besag–York–Mollié (BYM2) reparameterization, decomposing spatial variation into a structured component (*u*_*i*) capturing spatial dependence between neighboring counties and an unstructured independent and identically distributed (iid) component (*v*_*i*). specified as an independent and identically distributed (iid) random effect. The BYM2 specification uses a joint precision parameter and a mixing parameter *ɸ* ∈ [0,1], where higher values indicate greater residual spatial variance attributable to the structured component. Temporal trends were modeled using a first-order random walk (RW1) prior (*w_t*), and a space-time interaction term (*δ*_*it*) captures residual county-year variation as an iid effect.^26,27^

Model parameters were estimated via Integrated Nested Laplace Approximations (R-INLA package; v22.x) with penalized complexity priors. Fixed-effect posterior summaries were exponentiated to derive incidence rate ratios (IRRs) with corresponding 95% credible intervals (CrIs). Two models were estimated: (1) SREI as a continuous standardized score (IRR per 1-SD increase), and (2) SREI as quartiles (Q1-Q4, reference Q1), estimating IRR across Q2-Q4 relative to Q1. Both models adjusted for standardized z–scores of continuous covariates (education, worked full-time, married, insurance, and urban/rural status). Bayesian model assumptions were rigorously assessed for robustness, alternative priors, and potential nonlinearity (Supplementary file: Technical details). Posterior fitted values and predictive performances were mapped to visualize the spatial mortality risk pattern. Model fit was assessed using deviance, the Deviance Information Criterion (DIC), Watanabe–Akaike Information Criterion (WAIC), and conditional predictive ordinate (CPO), and deviance. ^26,27^

### Population Attributable Fraction (PAF) and Attributable Deaths (AD)

PAF represents the fraction of cases attributable to a risk factor; AD estimates the potentially preventable burden through intervention.^28,29^ PAF and AD were estimated under both continuous and categorical SERI specifications to quantify CVD mortality attributable to place-based neighbourhood disadvantage. AD was obtained by multiplying PAF by the total observed CVD deaths. For the continuous SREI, PAF was estimated via model-based counterfactual predictions,^28,30^ integrating full posterior uncertainty from the Bayesian joint distribution to ensure that all errors and geographic variations were accurately reflected. For continuous SREI, PAF was calculated as:

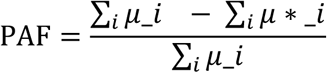

Where *μ*_*i* and *μ* ∗ _*i* are model-predicted and counterfactual predicted deaths, respectively. For categorical SREI,

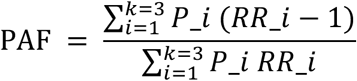

where *p*__*i*_ denotes the population proportion in the quartile *i* (*k* = 3; *Q*2 − *Q*4) and *RR*_*_i_* is the relative risk compared to Q1.^28,30^

### Sensitivity Analyses

Several sensitivity analyses evaluated model robustness. First, alternative continuous SREI parameterizations to assess potential nonlinearity. Second, varying spatial smoothing configurations evaluated sensitivity to spatial dependence assumptions. Third, alternative prior distributions for the spatial and temporal hyperparameters confirmed that findings were data-driven, rather than prior-driven. Finally, analyses were stratified by urban and rural counties separately to assess the consistency across geographic contexts.

## RESULTS

### Baseline characteristics

Baseline characteristics for 3,007 U.S. counties (2017-2020), stratified by SREI quartile, are displayed in Table 1. Rural counties increased from 52% in Q1 to 79% in Q4, reflecting the geographic convergence of structural disadvantages and rurality. The median proportion of Black residents was lower in Q1 (1% [0-5%]) than in Q4 (13% [2-35%]), supporting the SREI’s capture of racialized structural disadvantage. Compared with Q1, Q4 counties had significantly lower median household income ($64,460 versus $40,213), bachelor’s degree attainment (93% versus 82%), and homeownership (16,036 versus 7,271), but elevated disability rates (12% versus 20%) and social vulnerability (0.2 versus 0.8), indicating systematic sociodemographic inequities that may compound CVD mortality disparities in this demographic.

**Table 1:** Characteristics of the US counties (n = 3007), 2017 to 2020, stratified by Structural Racism Effect Index (SREI) Quartiles.

| Characteristics* | Overall<br>n = 3007 | Quartile 1<br>n = 767 | Quartile 2<br>n = 751 | Quartile 3<br>n = 744 | Quartile 4<br>n = 745 | P<br>value† |
| --- | --- | --- | --- | --- | --- | --- |
| **Percent Male,**<br>Median (IQR) | 45.4<br>(44.4 - 46.6) | 45.5<br>(44.7 - 46.6) | 45.8<br>(44.6 - 46.9) | 45.0<br>(44.0 - 46.2) | 44.70<br>(43.45 - 45.95) | <0.001 |
| **Percent Female,**<br>Median (IQR) | 54.6<br>(53.4 - 55.6) | 54.5<br>(53.4 - 55.3) | 54.2<br>(53.1 - 55.4) | 55.0<br>(53.8 - 56.0) | 55.30<br>(54.05 - 56.55) | <0.001 |
| **Percent Married,**<br>Median (IQR) | 64.0<br>(57.0 - 69.0) | 68.0<br>(63.0 - 73.0) | 64.0<br>(59.0 - 69.0) | 62.0<br>(57.0 - 67.0) | 58.00<br>(50.00 - 64.00) | <0.001 |
| **Percent ≥Bachelor**<br>**Education,**<br>Median (IQR) | 89.2<br>(84.7 - 92.2) | 93.3<br>(91.7 - 94.5) | 90.7<br>(88.8 - 92.3) | 87.6<br>(84.9 - 89.6) | 82.4<br>(79.7 - 85.4) | <0.001 |
| **Percent White**<br>Median (IQR) | 81.0<br>(63.0 - 90.0) | 85.0<br>(75.0 - 91.0) | 84.0<br>(72.0 - 91.0) | 79.0<br>(63.0 - 90.0) | 63.00<br>(49.00 - 84.00) | <0.001 |
| **Percent Black,**<br>Median (IQR) | 2.0<br>(1.0 - 10.0) | 1.0<br>(0 - 4.0) | 1.0<br>(0 - 5.0) | 3.0<br>(1.0 - 9.0) | 13.00<br>(2.00 - 35.00) | <0.001 |
| **Hispanic,**<br>Median (IQR) | 5.0<br>(2.0 - 10.0) | 5.0<br>(3.0 - 10.0) | 5.0<br>(2.0 - 11.0) | 5.0<br>(2.0 - 13.0) | 3.00<br>(2.00 - 8.00) | <0.001 |
| **Percent Asian,**<br>Median (IQR) | 0.6<br>(0.3 - 1.3) | 1.1<br>(0.4 - 3.2) | 0.6<br>(0.4 - 1.4) | 0.5<br>(0.3 - 1.1) | 0.41<br>(0.24 - 0.67) | <0.001 |
| **Worked Full time,**<br>Median (IQR) | 39.2<br>(38.4 - 40.1) | 39.0<br>(38.2 - 40.1) | 39.0<br>(38.3 - 39.8) | 39.2<br>(38.5 - 40.2) | 39.40<br>(38.70 - 40.20) | <0.001 |
| **Percent Living with**<br>**Disability,**<br>Median (IQR) | 15.6<br>(12.9 - 18.7) | 12.2<br>(10.5 - 14.0) | 14.9<br>(13.3 - 16.9) | 17.0<br>(14.9 - 19.3) | 19.5<br>(16.4 - 22.5) | <0.001 |
| **House owner,**<br>Median (IQR), count | 10,581<br>(4,605-27,968) | 16,035<br>(4,364 - 66,081) | 14,339<br>(5,567 - 37,940) | 10,732<br>(5,478 - 23,241) | 7,271<br>(4,034 - 12,599) | <0.001 |
| **Median Household**<br>**Income,**<br>Median (IQR), USD | 51,704<br>(44,169 - 59,742) | 64,460<br>(57,714 - 76,610) | 54,857<br>(51,080 - 60,097) | 48,809<br>(44,872 - 52,813) | 40,213<br>(36,004 - 44,522) | <0.001 |
| **Urban versus Rural, n (%)** |  |  |  |  |  | <0.001 |
| Rural | 1,913 (64.0%) | 400 (52.0) | 449 (60.0) | 478 (64.0) | 586 (79.0) |  |
| Urban | 1,094 (36%) | 367 (48.0) | 302 (40.0) | 266 (36.0) | 159 (21.0) |  |
| **SVI** ‡,<br>Median (IQR), score | 0.5<br>(0.3 - 0.8) | 0.2<br>(0.1 - 0.3) | 0.4<br>(0.3 - 0.6) | 0.6<br>(0.5 - 0.7) | 0.8<br>(0.7 - 0.9) | <0.001 |
| **SNAP** §<br>Median (IQR), counts | 1,485<br>(622 - 3,545) | 1,245<br>(285 - 4,599) | 1,698<br>(548 - 4,637) | 1,476<br>(750 - 3,489) | 1,473<br>(839 - 2,653) | 0.002 |

\*All continuous variables presented as median (Interquartile Range, IQR), categorical variable as n (%)
†P-values derived from the Kruskal-Wallis rank sum test for continuous variables, and Pearson's Chi-squared test for categorical variables.
‡SVI: Social Vulnerability Index, derived from the United States Department of Agriculture (USDA) Economic Research Service (ERS) Food Access Research Atlas.
§SNAP: Supplementary Nutritional Assistance Program, data derived from the Agency for Toxic Substances and Disease Registry (ATSDR) and the CDC.

### CVD Mortality burden

Approximately 2.79 million CVD deaths were observed among older U.S. adults during 2017-2020, with a median cumulative county-level death of 75 (IQR: 35-175). Table 2 summarizes observed and expected deaths, age-adjusted mortality rate (AAMR), and corresponding 95% confidence intervals, and dispersion matrices (variance and the coefficient of variation) across the SREI quartiles. AAMR increased progressively, from 1,290.8 per 100,000 in Q1 to 1,700.4 per 100,000 in Q4. One-way ANOVA demonstrated significant differences across quartiles (F = 845.6, p < 0.001), and Tukey HSD tests verified all pairwise comparisons (p <0.0001). Mortality variability also peaked in Q4 (Var: 141,244.20; CV: 0.2210), indicating greater mortality burden and geographic heterogeneity in counties with the highest structural disadvantage.

**Table 2:** Descriptive statistics of Deaths and Age-adjusted CVD mortality (AAMR) per 100,000, stratified by SREI Quartiles across 3007 counties.

| **SREI Quartile**<br>(n) | **Deaths**<br>Median (Min-Max) | **Total Death**<sup>*</sup><br>n (%) | **Expected Death**<br>(Min - Max) | **AAMR**<sup>†</sup><br>Mean (95% CI) | **Var**<sup>‡</sup> | **CV**<sup>§</sup> |
| --- | --- | --- | --- | --- | --- | --- |
| **Q1**<br>(2933) | 100 (10 – 19658) | 1128660 (40.5) | 4.01 - 18948.83 | 1290.90 (1015.90 - 1532.30) | 67795.17 | 0.20 |
| **Q2**<br>(2930) | 98 (10 – 8369) | 877411 (31.5) | 3.23 - 9573.74 | 1439.60 (1132.80 – 1727.30) | 75697.52 | 0.19 |
| **Q3**<br>(2930) | 82 (10 – 5263) | 515346 (18.5) | 6.02 - 4716.43 | 1555.40 (1219.90 - 1889.60) | 87978.08 | 0.19 |
| **Q4**<br>(2929) | 59 (10 – 1585) | 266735 (9.6) | 4.30 - 1435.63 | 1700.00 (1248.70 - 2128.90) | 141244.20 | 0.22 |

<sup>\*</sup>Total Death, n (%): (Sum deaths in quartile/Deaths across all data) \*100
<sup>†</sup>AAMR: Age-adjusted Mortality Rate of mortality of CVD per 100,000, using direct standardization on the 2000 US standard population;
<sup>‡</sup>Var: Variance of AAMR;
<sup>§</sup>CV: Coefficient of Variation of AAMR.
<sup>||</sup>Expected Death: Older adult Population \* ([sum Deaths in county] / [sum of older adult Population in the county])

### Exploratory analysis

AAMR per 100,000 demonstrated a marked geographic gradient across the US counties during the 2017–2020 period (Figure 1). The highest mortality rates (Q4: 1,692–3,598 per 100,000) clustered in the Deep South, Mississippi Delta, Appalachia, Oklahoma, and the Desert Southwest. In contrast, the lowest rates (Q1: 508–1,268 per 100,000) were observed in the Northern Plains, Upper Midwest, and parts of the Northeast. Data suppression (represented by white space) was most common in the Western U.S. and the Great Plains because of low death counts. SREI scores also showed marked regional clustering (median: 42 [IQR: –0.05 to 0.91]), with the highest structural disadvantage concentrated across the Southeast, Mississippi Delta, and the historic Black Belt, whereas lower SREI quartile counties predominated in the Upper Midwest, Northern Plains, and northern New England (Figure 2). Distribution of AAMR and expected deaths by SREI quartiles is displayed in Figure S1.

**Figure 1.**
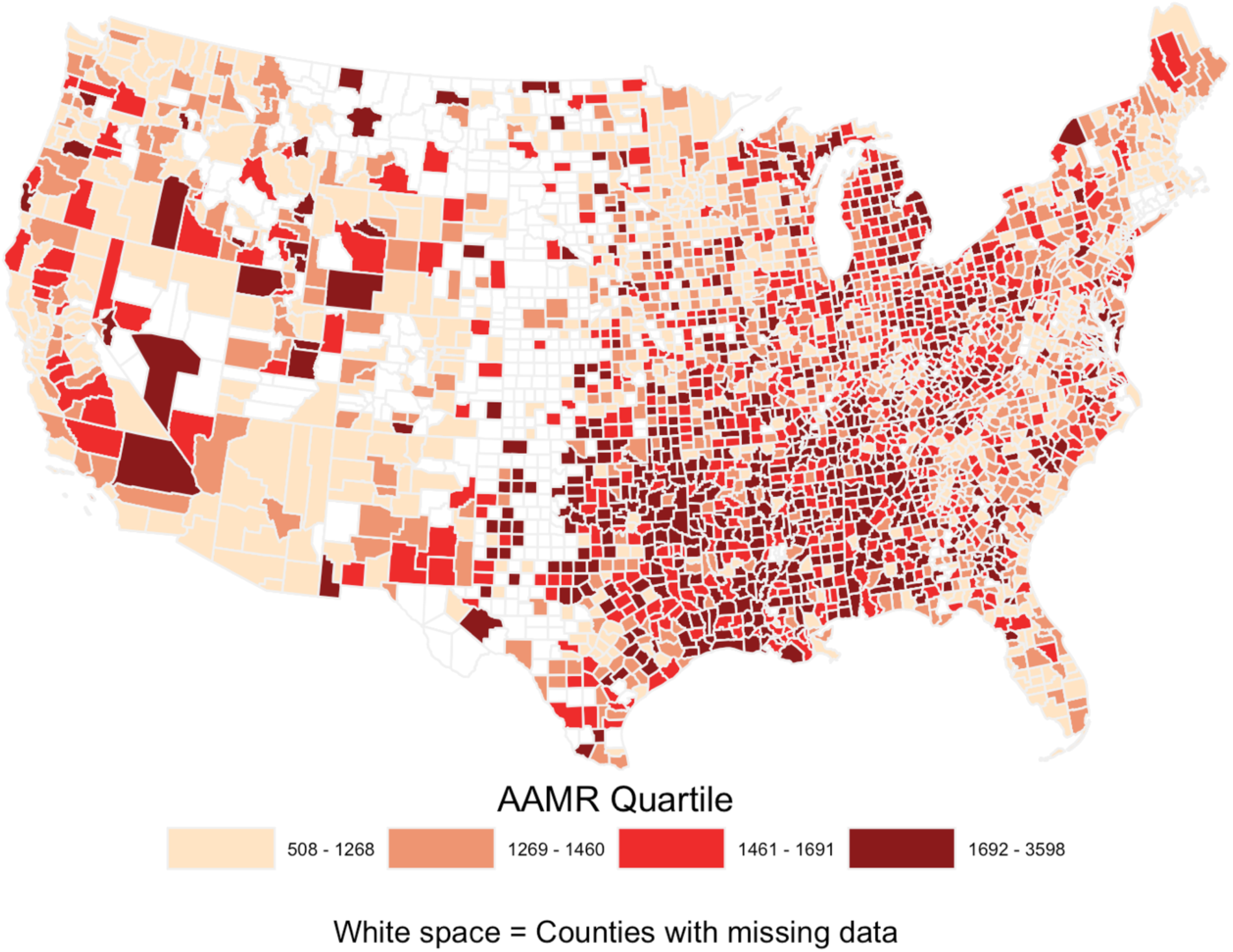
Age-Adjusted Mortality Rate per 100,000 during 2017 to 2020 among older US adults. Choropleth maps illustrate the spatiotemporal distribution of Age-adjusted Mortality rate (AAMR) per 100,000 by US county, during 2017–2020. White space indicates counties excluded due to CDC WONDER data suppression thresholds. The highest mortality rates (Q4: 1,692–3,598 per 100,000) clustered in the Deep South, the Mississippi Delta, Appalachia, Oklahoma, and the Desert Southwest.

**Figure 2.**
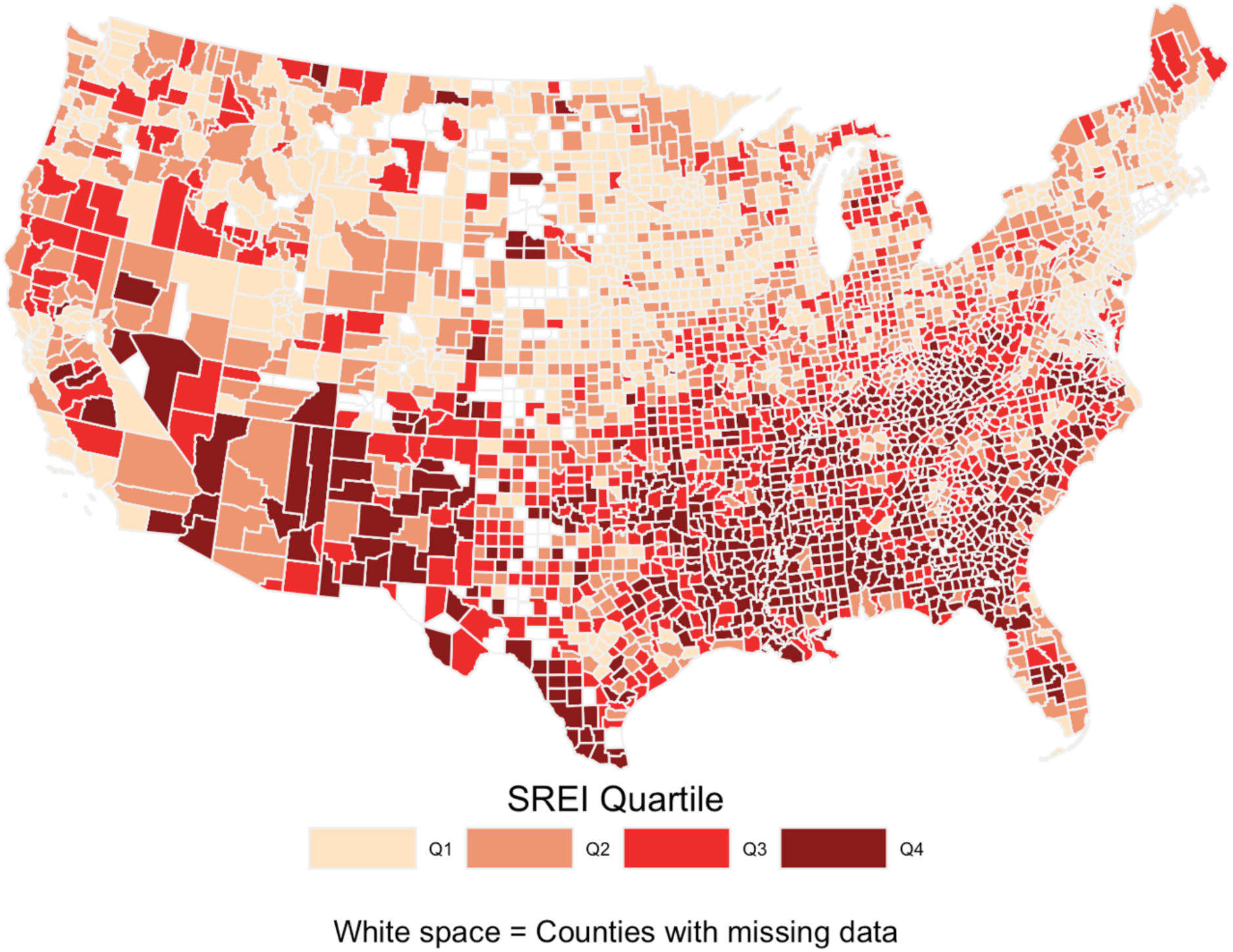
Geographic distribution of the Structural Racism Effect Index (SREI) by U.S. county. The choropleth map illustrates SREI quartiles across the United States, with darker shading indicating a higher burden of structural racism. Spatial clustering of the fourth quartile (Q4) is evident in the Southeast, the Mississippi Delta, and the historic Black Belt.

### Spatial Autocorrelation

Global Moran’s I exhibited significant positive spatial autocorrelation in AAMRs (I = 0.35, 95% CI: 0.32 to 0.37; p < 0.001), indicating that neighboring counties had similar mortality rates. Figure 3 displayed High-High hotspots in the Southeast and Low-Low cold spots across the Great Plains, Upper Midwest, and Mountain West, with High-Low and Low-High outliers highlighting localized departures from surrounding mortality patterns.

**Figure 3.**
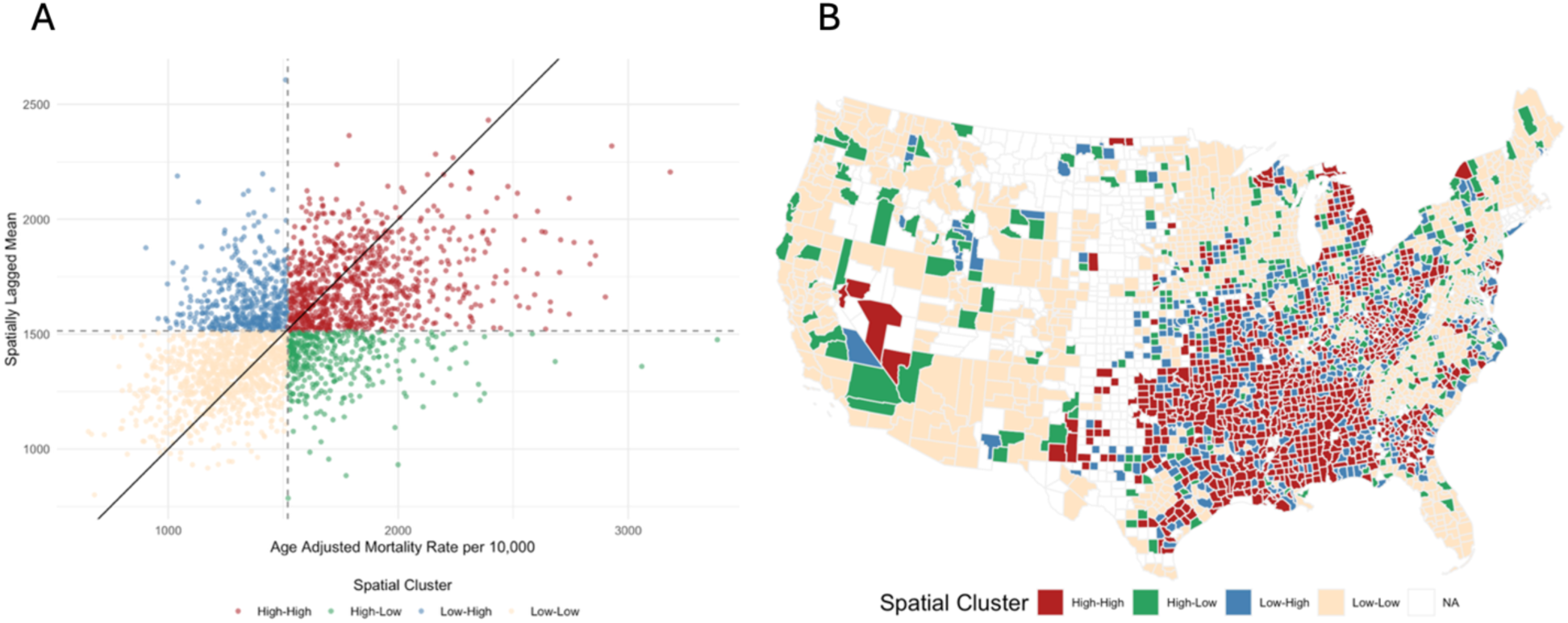
Moran scatterplot of age-adjusted CVD mortality (A), and Moran’s I Cluster Map of AAMR across U.S. counties (B). The scatterplot and map visualize spatial autocorrelation by comparing local mortality rates against spatially lagged means, with quadrants identifying clusters of similar values: High-High (Red), Low-Low (Beige), High-Low (Green), and Low-High (Blue). The cluster map highlights a prominent geographic concentration of high mortality rates (High-High clusters) across the southeastern United States, contrasted with lower mortality clusters (Low-Low) scattered predominantly throughout the northern and western regions.

### Association of SREI and Cardiovascular Mortality

In the fully adjusted continuous model, a 1-SD increase in the SREI was associated with 13% higher risk of CVD mortality (IRR: 1.13, 95% CrI: 1.12–1.15, *p* <0.001), independent of sociodemographic covariates including education, employment, marital status, insurance coverage, and urban/rural classification. The quartile-based model revealed a monotonic gradient, with progressively higher risk in Q2 (IRR: 1.10, 95% CrI: 1.08–1.12), Q3 (IRR: 1.16, 95% CrI: 1.13–1.19), and Q4 (IRR: 1.24, 95% CrI: 1.20 –1.28), compared to Q1, when adjusted for the same covariates. Among covariates, county-level full-time employment was positively associated with CVD mortality (IRR: 1.02, 95% CrI: 1.01–1.03) across models. Model estimates are displayed in Figure 4. The BYM2 spatial hyperparameters indicated most residual spatial variation was structured (*φ* = 0.83, 95% CrI: 0.77–0.88). Temporal variability was low, with high precision in the random-walk component (τ ≈ 2800–2910). Model fit statistics (Table S2) favored the adjusted space-time interaction framework (BYM2+RW1+ iid S*T), which achieved the lowest DIC (90632.36), WAIC (90814.49), and deviance (85817.69), and favorable CPO (–3.90). Sensitivity analyses were materially unchanged, supporting the robustness of the associations (Table S3). In stratified analyses, the SREI-CVD mortality association was IRR: 1.14 (95% CrI: 1.12–1.17) among urban counties and IRR: 1.11 (95% CrI: 1.09–1.13) among rural counties. The exposure-response relationship between SREI and posterior relative risk of CVD deaths is displayed in Figure S2. Spatial distribution of posterior mean fitted value and posterior uncertainty is displayed in Figure S3-4.

**Figure 4.**
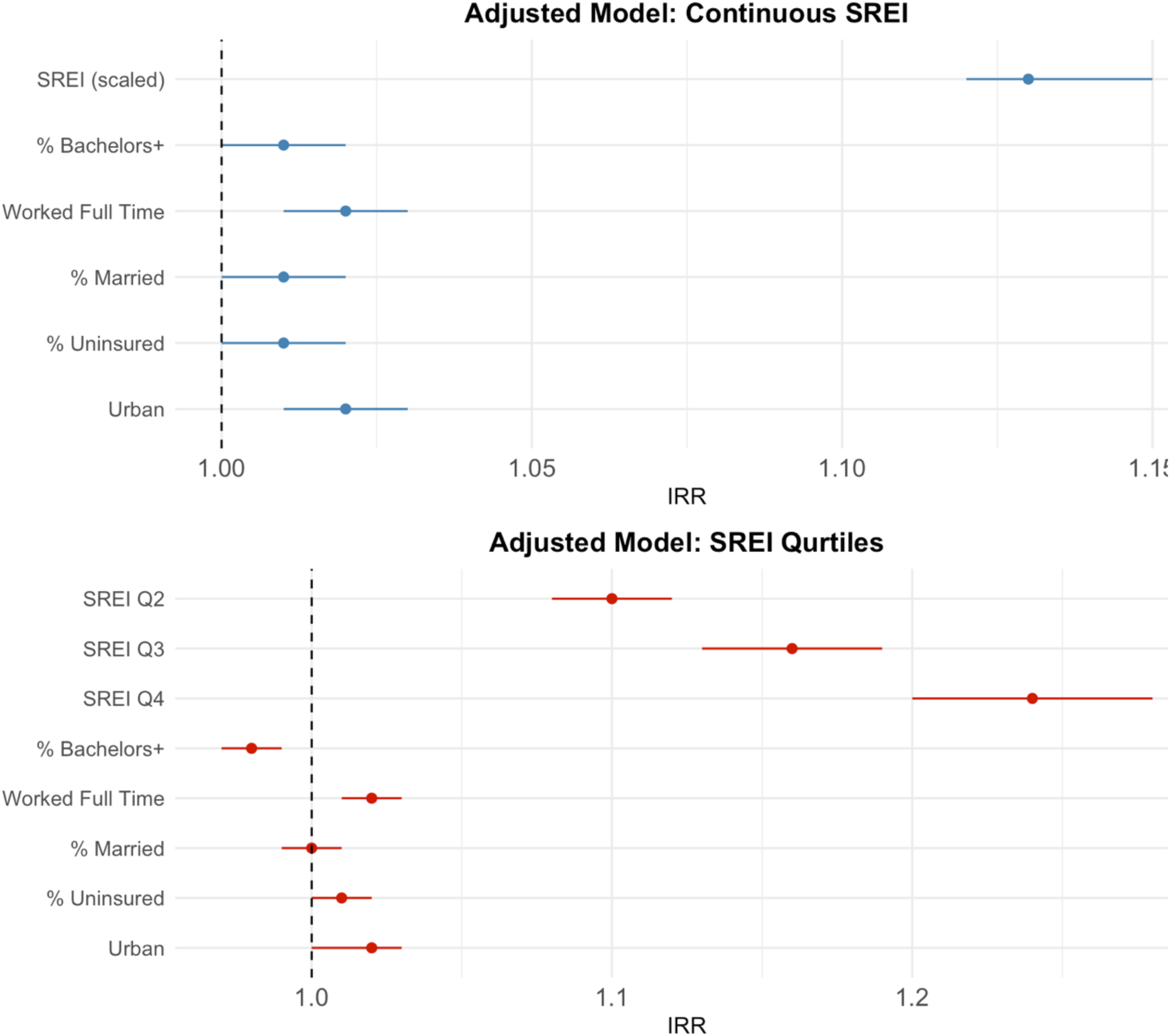
Association between the Structural Racism Effect Index (SREI) and CVD mortality. The estimates represent incidence risk ratios (IRRs) derived from both continuous (standardized SREI) and quartile-based multivariable Bayesian spatiotemporal models, adjusted for county-level sociodemographic characteristics, including educational attainment, full-time employment status, marital status, insurance coverage, and urban-rural classification.

### Population Attribution Fraction (PAF) and Attributable Deaths (AD)

Among adults aged ≥65 years, the PAF and AD associated with higher SREI are summarized in Table 3. In the continuous model, higher SREI accounted for 6.94% of CVD deaths (95% CI: 6.13%–7.73%), corresponding to 193,472 potentially preventable deaths (95% CI: 171,052–215,537) over the study period, under a counterfactual scenario of eliminating neighborhood disadvantage. In the quartile-based model, PAF demonstrated a monotonic increase relative to Q1, rising from 2.12% (95% CI: 1.70 – 2.56) in Q2, 3.50% (95% CI: 2.95 – 4.06) in Q3, and 5.23% (95% CI: 4.45 – 6.03) in Q4. In contrast, AD exhibited a reverse trend, the highest burden observed in Q2 (18,641 deaths, 95% CI: 14893 – 22446), versus the lowest burden in Q4 (13,962 deaths, 95% CI: 11,870 – 16,077). This divergence indicates that while relative risk gradients increase with deprivation, the absolute burden is additionally shaped by population distribution across exposure strata.

**Table 3:** Estimated Population Attributable Fraction (PAF) and Attributable Deaths (AD) among the 65+ population.

| **Predictors** | **PAF, % (95% CI)** | **Attributable death (95%CI)\*** |
| --- | --- | --- |
| **SREI** (Continuous scale) | 6.94 (6.13 – 7.73) | 193,472 (171,052 – 215,537) |
| **SREI Quartile** (Reference = Quartile 1) |  |  |
| Quartile 2 | 2.12 (1.70 – 2.56) | 18,641 (14,893 – 22,446) |
| Quartile 3 | 3.50 (2.95 – 4.06) | 18,053 (15,209 – 20,944) |
| Quartile 4 | 5.23 (4.45 – 6.03) | 13,963 (11,870 - 16,077) |

\*Attributable death presented in the nearest rounded value.

### Spatial Exceedance Probability

Spatial exceedance probability analysis (Figure 5) revealed marked geographic variation in excess CVD mortality risk. Counties with the high posterior probabilities of elevated risk (>0.95) clustered predominantly in the Southeast and parts of the Southwest, indicating strong evidence of mortality exceeding expected baseline levels. In contrast, low exceedance probabilities (<0.20) were concentrated across the Great Plains, Upper Midwest, and Mountain regions.

**Figure 5.**
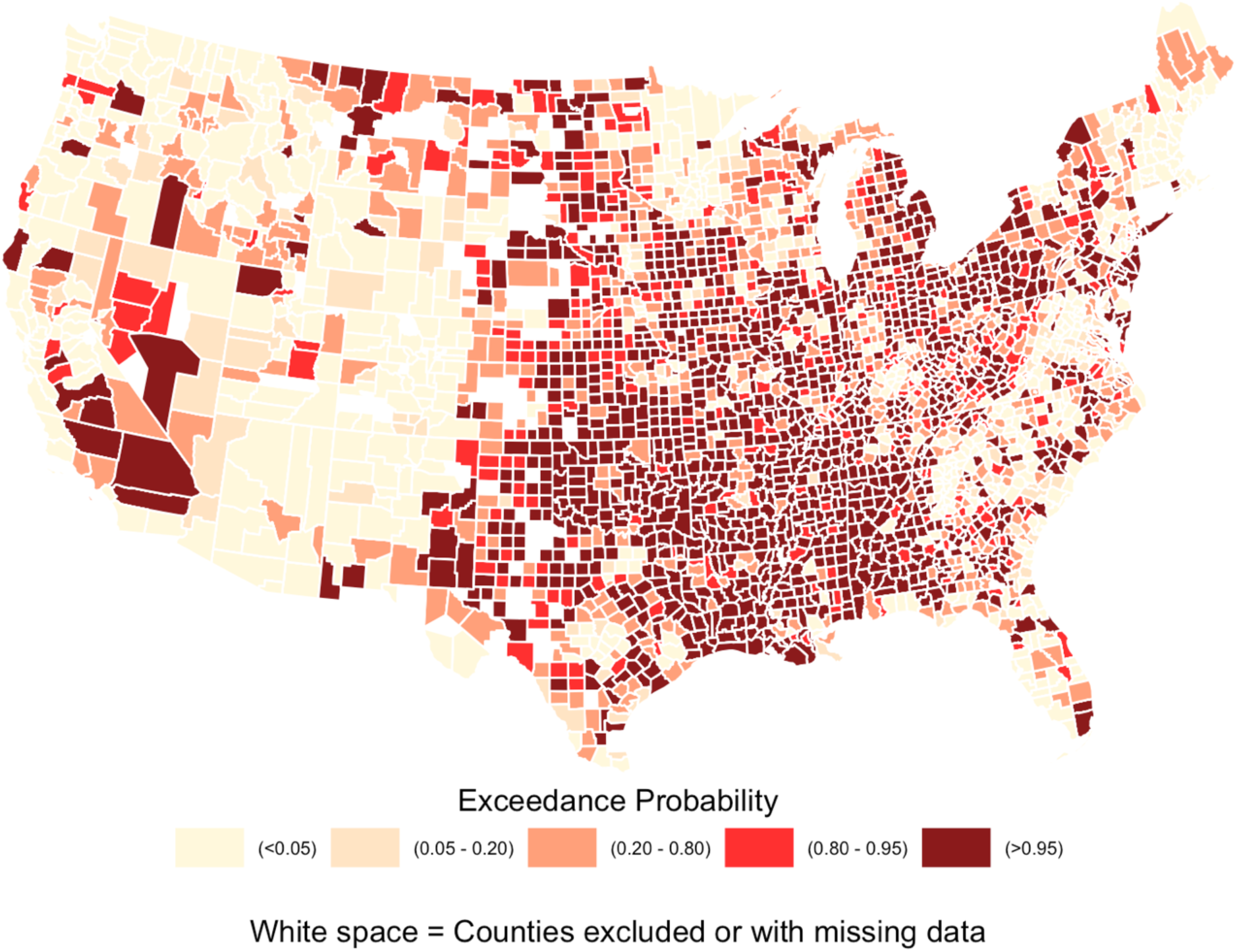
Posterior exceedance probability (RR >1) of CVD mortality exceeding the national expected rate by U.S. county. High exceedance probabilities (P >0.95) are heavily clustered throughout the Southeast, Midwest, and parts of the Southwest, indicating areas where the death rate almost certainly exceeds the national average.

## DISCUSSION

This Bayesian spatiotemporal ecological study of 3,007 U.S. counties (2017-2020) demonstrated neighborhood structural racism as a significant, dose-dependent predictor of CVD mortality among adults aged ≥65 years. Each 1-SD increase in the SREI was associated with a 13% increase in CVD mortality risk, with a monotonic gradient from 10% (Q2) to 24% (Q4), relative to Q1. At the population level, a counterfactual reduction of SREI to average levels could theoretically prevent ∼7% of excess CVD mortality, translating to 193,472 potentially avoidable deaths over the study period. Crucially, categorical absolute burden analysis revealed an inverse burden gradient: the absolute number of avoidable deaths (AD) peaked in mildly disadvantaged counties (Q2: 18,641 deaths) rather than the most extreme enclaves (Q4: 13,963 deaths), exposing a critical nuance for structural intervention design.

These findings extend prior evidence linking structural racism as a significant predictor of CVD deaths among older adults, underscoring the potential to avoid this burden through population-level structural interventions. Safford et al. demonstrated an association between state-level structural factors and incident coronary heart disease,^13^ while Lawrence et al. and Liu et al. identified neighbourhood-level inequities through composite indices;^16,17^ however, these studies focused on intermediate cardiovascular health metrics rather than hard mortality endpoints. The observed dose-response gradient supports the theoretical frameworks positing that structural racism operates through cumulative, multi-level pathways—restricted healthcare access, concentrated poverty, environmental toxicity, and chronic psychosocial stress—to amplify cardiovascular risk over the life course.^7^ This graded pattern (24 % increased risk of mortality in Q4 counties, compared to Q1) corroborates progressive associations between social vulnerability and clinical CVD reported by Ibrahim et al. and Bevan et al.^4,14^ Methodologically, the significant Moran’s I (I = 0.35; p <0.001) reveals spatial structuring of CVD mortality, reflecting shared healthcare infrastructures, policy environments, and institutional legacies across neighboring countries.^31^ Notably, 83% of residual spatial heterogeneity was attributable to structured geographic clustering (BYM2 φ=0.83), with the highest exceedance probabilities (>0.95) concentrated in the Southeast, the Mississippi Delta, and Southwest – regions overlapping the well-documented “Stroke Belt”,^32^ and historic redlining contours.^7,8^ The exceptionally high temporal random walk precision (mean≈2900) further indicates these geographic inequities remained deeply stable throughout the 2017–2020, underscoring the inertial persistence of structural disadvantage and limited penetration of equity-focused policies during this interval.^18^ From a public health perspective, a ∼7% PAF demonstrates that the population burden of structural racism is a modifiable determinant comparable in magnitude to routinely targeted clinical and behavioral risk factors.^3,28,30^ Crucially, our absolute burden exposes a manifestation of Rose’s prevention paradox: the largest attributable death burden occurred in Q2 (18,641 deaths) rather than Q4 (13,963 deaths), as mildly disadvantaged counties collectively encompass a far larger at-risk population.^30,33^ Given that older adults account for ∼80% of national CVD fatalities,^2^ this population warrants priority for upstream intervention. Policies narrowly targeting only the most disadvantaged communities will systematically miss the greater aggregate mortality burden distributed across mildly to moderately disadvantaged areas. Maximizing impact requires broad, population-wide investments in housing equity, educational access, and economic opportunity that shift the entire risk distribution downward.^3,30^ Prioritizing such reforms in historically burdened regions could prevent an estimated 193,472 deaths while lowering average risk across the population.

This study has notable strengths. The multidimensional SREI captures compounding structural exposures rather than proxy endpoints. The Bayesian spatiotemporal framework simultaneously addresses spatial autocorrelation and temporal dynamics, yielding more precise estimates than cross-sectional approaches. Integrating both continuous and categorical exposures reveals a monotonic relative risk gradient while simultaneously exposing the reverse trend of absolute public health burdens because of the underlying population distribution. Finally, translating these into PAF and AD metrics provides policy-relevant quantification of potentially preventable mortality. aligning with Rose’s prevention paradox. However, several limitations warrant consideration. The ecological design precludes individual-level causal inferences. Despite comprehensive covariate adjustment, residual confounding from unmeasured behavioral (e.g., smoking), environmental (e.g., pollution), or health system factors (e.g., migration, local clinical quality) may persist. Additionally, reliance on CDC WONDER death certificate coding introduces potential outcome misclassification, and the exclusion of island territories could modestly limit broader national generalizability.

In conclusion, neighborhood structural racism represents a spatially structured, dose-dependent predictor of CVD mortality among older US adults. A substantial proportion of this mortality burden appears attributable to modifiable structural conditions, reinforcing the imperative to integrate place-based determinants into CVD prevention frameworks alongside traditional biological and behavioral targets. These geographically granular findings can guide resource allocation and policy prioritization. Future individual-level research disaggregating SREI components is needed to identify which specific conditions most contribute to these inequities and to inform precision policy interventions.

## DECLARATIONS

### Data Availability Statement

The data presented in this study were derived from publicly available deidentified data, derived from resources provided by the <u>CDC</u>, accessed on April 13, 2026. Exposure data was collected from the <u>SREI database</u> on April 12, 2026. American Community Survey data was collected from the <u>US Census Bureau</u> database on April 13, 2026. Supplemental Nutrition Assistance Program (SNAP) participation from the <u>United States Department of Agriculture (USDA) Economic Research Service (ERS) Food Access Research Atlas</u>. The Social Vulnerability Index (SVI) was developed by the <u>Agency for Toxic Substances and Disease Registry (ATSDR) and the CDC database</u> on April 13, 2006.

## Acknowledgements

We gratefully acknowledge CDC WONDER, the SREI database, the American Community Survey (ACS), the USDA, and the CDC/ATSDR, as well as the individuals who participated in these datasets and made this research possible. We also express our sincere gratitude to Dr. Rajib Paul, Professor of Epidemiology and Community Health at the University of North Carolina at Charlotte, USA, for his invaluable mentorship in spatial epidemiology.

## Statistical Code Availability Statement

Code available from TB upon reasonable request

## Ethics approval

The datasets are publicly accessible and de-identified, and therefore, it is exempt from Institutional Review Board (IRB) approval.

## Consent for publication

Not applicable.

## CRediT Authorship contribution statement

**TB:** Conceptualization and design, data acquisition, formal statistical analysis and interpretation of results, drafting manuscript, reviewing critically for intellectual content, and editing. **MS**: Revised critically for intellectual content and editing. **HC**: Revised critically for intellectual content and editing. All authors read and approved the final manuscript.

## Statement on AI use

During the preparation of this work, the authors used Claude (Version Sonnet 4.6) and Google Gemini (3.5 Flash) to improve the readability of the manuscript. After using this tool, the authors reviewed and edited the content as needed and take full responsibility for the content of the publication.

## Non-standard Abbreviations and Acronyms

ACS: American Community Survey
AHA: American Heart Association
AD: Attributable Death
AR1: First-Order Autoregressive
AAMR: Age-Adjusted Mortality Rate
BYM: Besag–York–Mollié
CDC: Centers for Disease Control and Prevention
CPO: Conditional Predictive Ordinate
CrI: Credible Interval
CV: Coefficient of Variation
CVD: Cardiovascular disease
DIC: Deviance Information Criterion
FIPS: Federal Information Processing Standard
ICD-10: International Classification of Disease-10
IID: Independent and Identically Distributed
INLA: Integrated Nested Laplace Approximations
IQR: Interquartile Range
IRR: Incidence Risk Ratio
PAF: Population Attribution Factor
RR: Relative Risk
RW1: First-order Random Walk
SNAP: Supplemental Nutrition Assistance Program
SREI: Structural Racism Effect Index
SVI: Social Vulnerability Index
WAIC: Watanabe-Akaike Information Criterion
WONDER: Wide-Ranging Online Data for Epidemiologic Research

## REFERENCES

1. Palaniappan LP, Allen NB, Almarzooq ZI, Anderson CAM, Arora P, Avery CL, Baker-Smith CM, Bansal N, Currie ME, Earlie RS, Fan W, Fetterman JL, Gibbs BB, Heard DG, Hiremath S, Hong H, Hyacinth HI, Ibeh C, Jiang T, Johansen MC, Kazi DS, Ko D, Kwan TW, Leppert MH, Li Y, Magnani JW, Martin SS, Michos ED, Mussolino ME, Ogungbe O, Parikh NI, Perez M V., Perman SM, Sarraju A, Shah NS, Springer M V., St-Onge M-P, Thacker EL, Tierney S, Urbut SM, Spall HGC Van, Voeks JH, Whelton SP, Khan MSS. 2026 Heart Disease and Stroke Statistics: A Report of US and Global Data From the American Heart Association. Circulation. 2026;153:1–632.

2. Yazdanyar A, Newman AB. The Burden of Cardiovascular Disease in the Elderly: Morbidity, Mortality, and Costs. Clin. Geriatr. Med. 2009;25:563–577.

3. Magnussen C, Ojeda FM, Leong DP, Alegre-Diaz J, Amouyel P, Aviles-Santa L, Bacquer DDB, Ballantyne CM, Bernabé-Ortiz A, Bobak MB, Brenner H, Carrillo-Larco RM, Lemos J de, Dobson A, Dörr M, Donfrancesco C, Drygas W, Dullaart RP, Engström G, Ferrario MM, Ferrières JF, Gaetano G de, Goldbourt UG, Gonzalez C, Grassi G, Hodge AM, Hveem KH, Iacoviello L, Ikram MK, Irazola V, Jobe M, Jousilahti P, Kaleebu P, Kavousi M, Kee F, Khalili D, Koenig W, Kontsevaya A, Kuulasmaa K, Lackner KJ, Leistner DM, Lind L, Linneberg A, Lorenz T, Lyngbakken MN, Malekzadeh R, Malyutina S, Mathiesen EB, Melander O, Metspalu A, Miranda JJ, Moitry M, Nalini M, Nambi V, Ninomiya T, Oppermann K, d’Orsi E, Pająk A, Palmieri L, Panagiotakos D, Perianayagam A, Peters A, Poustchi H, Prentice AM, Prescott EP, Risérus U, Salomaa V, Sans S, Sakata S, Schöttker B, Schutte AE, Sepanlou SG, Sharma SK, Shaw JE, Simons LA, Söderberg S, Tamosiunas A, Thorand B, Twerenbold R, Vanuzzo D, Veronesi G, Waibel J, Wannamethee SG, Watanabe M, Wild PS, Yao Y, Zeng Y, Ziegler A, Blankenberg S. Global Effect of Modifiable Risk Factors on Cardiovascular Disease and Mortality. New England Journal of Medicine. 2023;389:1273–1285.

4. Bevan G, Pandey A, Griggs S, Dalton JE, Zidar D, Patel S, Khan SU, Nasir K, Rajagopalan S, Al-Kindi S. Neighborhood-level Social Vulnerability and Prevalence of Cardiovascular Risk Factors and Coronary Heart Disease. Curr. Probl. Cardiol. 2023;48. doi:10.1016/j.cpcardiol.2022.101182.

5. Kundrick J, Rollins H, Mullachery P, Sharaf A, Schnake-Mahl A, Diez Roux A V., Bilal U. Heterogeneity in disparities by income in cardiovascular risk factors across 209 US metropolitan areas. Prev Med Rep. 2024;47. doi:10.1016/j.pmedr.2024.102908.

6. An DH. CVD Mortality Disparities with Risk Factor Associations Across U.S. Counties. Healthcare (Switzerland). 2025;13. doi:10.3390/healthcare13222937.

7. Khadke S, Kumar A, Al-Kindi S, Rajagopalan S, Kong Y, Nasir K, Ahmad J, Adamkiewicz G, Delaney S, Nohria A, Dani SS, Ganatra S. Association of Environmental Injustice and Cardiovascular Diseases and Risk Factors in the United States. Journal of the American Heart Association. 2024;13. doi:10.1161/JAHA.123.033428.

8. Kraus NT, Connor S, Shoda K, Moore SE, Irani E. Historic redlining and health outcomes: A systematic review. Public Health Nurs. 2024;41:287–296.

9. Williams DR, Lawrence JA, Davis BA. Racism and Health: Evidence and Needed Research. Annu. Rev. Public Health.. 2019;40:105–125.

10. Diez Roux A V., Mair C. Neighborhoods and health. Ann. N. Y. Acad. Sci.. 2010;1186:125–145.

11. Albert MA, Churchwell K, Desai N, Johnson JC, Johnson MN, Khera A, Mieres JH, Rodriguez F, Velarde G, Williams DR, Wu JC. Addressing Structural Racism Through Public Policy Advocacy: A Policy Statement From the American Heart Association. Circulation. 2024;149:E312–E329.

12. Bailey ZD, Krieger N, Agénor M, Graves J, Linos N, Bassett MT. Structural racism and health inequities in the USA: evidence and interventions. The Lancet. 2017;389:1453–1463.

13. Safford MM, Brown T, Bryan J, Brown TM, Pinheiro L. State-Level Structural Racism and Incident Coronary Heart Disease. J Am Heart Assoc. 2026;15:e039828.

14. Ibrahim BB, Barcelona V, Condon EM, Crusto CA, Taylor JY. The Association between Neighborhood Social Vulnerability and Cardiovascular Health Risk among Black/African American Women in the InterGEN Study. Nurs Res. 2021;70:S3–S12.

15. Fisher V, Boretsky A, Alkhouri N, Abuelezam NN. Then and now: umbrella variables mask temporal mechanisms of structural racism in US urban environments. Crit Public Health. 2025;35. doi:10.1080/09581596.2025.2492798.

16. Liu M, Patel VR, Salas RN, Rice MB, Kazi DS, Zheng ZN, Wadhera RK. Neighborhood Environmental Burden and Cardiovascular Health in the US. JAMA Cardiol. 2024;9:153–163.

17. Lawrence WR, Hong HG, Williams F, Dyer Z, Bergeron NQ, Brewer LPC, Chen Y, Crittendon DR, Freedman ND, Haas CB, Jackson SS, Martz CD, Mcgee-Avila JK, Ormiston CK, Pichardo CM, Rogers CR, Santiago-Rodríguez EJ, Shariff-Marco S, Turney IC, Powell-Wiley TM, Zhang W, Shiels MS. Manifestations of Structural Racism and Inequities in Cardiovascular Health Across US Neighborhoods. JAMA Health Forum. 2025;6. doi:10.1001/jamahealthforum.2025.3864.

18. Meliker JR, Sloan CD. Spatio-temporal epidemiology: Principles and opportunities. Spat. Spatiotemporal Epidemiol. 2011;2:1–9.

19. Friede A, Reid JA, Ory HW. CDC WONDER: A Comprehensive On-Line Public Health Information System of the Centers for Disease Control and Prevention.

20. Dyer Z, Alcusky MJ, Galea S, Ash A. Measuring The Enduring Imprint Of Structural Racism On American Neighborhoods. Health Aff. 2023;42:1374–1382.

21. ACS. DATA FREQUENTLY ASKED QUESTIONS Community Development Financial Institutions (CDFI) Fund New Markets Tax Credit (NMTC) Low-Income Communities (LIC).; 2023.

22. You W, Zhang G, Davy BM, Carlson A, Lin BH. Food consumed away from home can be a part of a healthy and affordable diet. Journal of Nutrition. 2009;139:1994–1999.

23. Flanagan BE, Gregory EW, Hallisey EJ, Heitgerd JL, Lewis B. A Social Vulnerability Index for Disaster Management. J Homel Secur Emerg Manag. 2020;8. doi:10.2202/1547-7355.1792.

24. von Elm E, Altman DG, Egger M, Pocock SJ, Gøtzsche PC, Vandenbroucke JP. The Strengthening the Reporting of Observational Studies in Epidemiology (STROBE) statement: guidelines for reporting observational studies. J Clin Epidemiol. 2008;61:344–349.

25. Anderson RN, Rosenberg HM. Age Standardization of Death Rates: Implementation of the Year 2000 Standard.

26. Rue H, Martino S, Chopin N. Approximate Bayesian inference for latent Gaussian models by using integrated nested Laplace approximations.; 2009: 1–319–392.

27. Blangiardo M, Cameletti M, Baio G, Rue H. Spatial and spatio-temporal models with R-INLA. Spat. Spatiotemporal Epidemiol. 2013;4:33–49.

28. Steenland K, Armstrong B. An overview of methods for calculating the burden of disease due to specific risk factors. Epidemiology. 2006;17:512–519.

29. Putrik P, Otavova M, Faes C, Devleesschauwer B. Variation in smoking attributable all-cause mortality across municipalities in Belgium, 2018: application of a Bayesian approach for small area estimations. BMC Public Health. 2022;22. doi:10.1186/s12889-022-14067-y.

30. Rockhill B, Newman B, Weinberg C. Use and Misuse of Population Attributable Fractions. Am J Public Health. 1998;88:15–19.

31. Kramer MR, Waller LA. From Spatial Analysis to Spatial Thinking: Reframing the Role of Place in Epidemiology. Curr Epidemiol Rep. 2026;13:16.

32. Howard G, Howard VJ. Twenty Years of Progress Toward Understanding the Stroke Belt. Stroke. 2020;51:742–750.

33. Geoffrey R. Strategy of prevention: Lessons from cardiovascular disease. Br Med J (Clin Res Ed). 1981;282:1847–1851.

